# Diabetics in the United Arab Emirates at higher risk of foot ulceration-Clinical Implications from Kinetics at Ankle Joint

**DOI:** 10.1101/2024.02.18.24303003

**Authors:** Animesh Hazari, Praveen Kumar, Shashi Kumar

## Abstract

**Introduction:** The prevalence of diabetes mellitus in the United Arab Emirates stands significantly high. Considering the etiopathogenesis for diabetic foot ulcers, studies suggest that increased plantar pressure, also referred to as the peak plantar pressure beyond a threshold value leads to the breakdown of skin and thus causes an ulcer. The accurate analysis of ground reaction force and peak plantar pressure could be suggestive and predictive for the occurrence of foot ulceration among diabetes mellitus and the study aims to analyze the kinetics at the ankle joint among diabetics in the UAE.

**Methodology:** The cross-sectional study was conducted at the Thumbay Physical Therapy and Rehabilitation Hospital, Gulf Medical University, Ajman, United Arab Emirates. 38 out of a total of 120 participants screened and diagnosed with Type 2 diabetes mellitus were taken for data collection and analysis. BTS motion analysis and Wintrack foot scan mat were used for kinetics at the ankle joint.

**Results:** Higher mean peaks of plantar pressure, 810, 654, and 911 kPa respectively were found at all three phases of the gait cycle. A positive and high correlation was found between peak plantar pressure risk of foot ulcer (r= 0.84). A negative and moderate correlation was observed for ankle joint moment and power.

**Conclusion:** The diabetic population in the United Arab Emirates could be at higher risk of foot ulceration with significantly increased peak plantar pressure. The altered kinetics such as reduced ankle joint moment and power could further add to the risk.

## Introduction

Foot ulceration among diabetics is one of the most serious complications that affects all domains of life significantly. The fate of unhealed foot ulceration is amputation leading to a poor Quality of Life (QoL) (Wang et al. 2022). Previous studies suggest that foot ulceration among diabetics is associated with the clinical term “Diabetic Foot” which is a triad of peripheral neuropathy, vascular and muscular dysfunction (Volmer et al. 2016). The risk of ulceration increases with long-standing hyperglycemia, severity of neuropathy, and altered biomechanics (Miranda et al. 2021).

The prevalence of diabetes mellitus in the United Arab Emirates stands significantly high. In its 2021 report, the international diabetes federation (IDF) suggested a prevalence rate of 16.4% which is expected to increase to 18.1 in 2045 (IDF Atlas 2021). In terms of number of people suffering from the disease, a suggestive figure of 300 million has been predicted for the upcoming year 2025 (Shieb et al. 2020). Considering the etiopathogenesis for diabetic foot ulcers, studies suggest that increased plantar pressure, also referred to as the peak plantar pressure beyond a threshold value leads to the breakdown of skin and thus causes an ulcer (Armstrong et al. 1998). In the presence of Diabetic Peripheral Neuropathy (DPN), and associated altered/ absent sensory input, the excessive plantar pressure initially causes a callus formation which on repetitive pressure leads to unhealed ulceration. However, the occurrence of ulceration is biomechanically determinative (Bus et al. 2002). In comparison to a healthy population, those suffering from diabetes mellitus have higher chances of risk of foot ulceration even with normal ground walking. At different phases of the gait cycle, only part of the foot contacts the ground which could lead to increased peak pressure on that area, and thus the possibility of skin breakdown to ulcer formation is higher. People with diabetes mellitus often present with ulcers at the heel or metatarsal heads. These areas are prone to increased pressure at heel strike and toe-off phases of the gait cycle. The plantar pressure proportionately increases with other activities that include higher ground reaction forces such as jogging, running, jumping, squatting, etc. which further hinders the activities of the diabetic population. Thus, the accurate analysis of ground reaction force and peak plantar pressure could be suggestive and predictive for the occurrence of foot ulceration among diabetes mellitus (Hazari et al. 2019) and the study aims to analyze the kinetics at the ankle joint among diabetics in the UAE under the following defined objectives:

1. To analyze the peak plantar pressure, moment, and power at the ankle joint among Type 2 diabetes mellitus in the UAE
2. To determine the association of ankle joint kinetics to the risk of foot ulceration among diabetics in UAE.

## Methodology

A cross-sectional study was conducted at the Thumbay Physical Therapy and Rehabilitation Hospital, Gulf Medical University, Ajman, United Arab Emirates. Following ethical approval (IRB/COHS/FAC/21/May 2021), a written informed consent was obtained from all eligible participants recruited in the study under the purposive sampling method. The study was conducted from September 2021 to October 2023. A total of 120 participants were screened, out of which 38 diagnosed cases of Type 2 diabetes mellitus participants (based on the recent clinical biochemistry and physician diagnosis) were taken for data collection and analysis. The BTS system with 12 infrared cameras and a force plate was used to collect the kinetic data. Helen Hayes marker sets were used to define the foot model for static and dynamic gait analysis. The peak plantar pressure was collected at the heel strike, midstance, and toe-off phases of the gait cycle using the Win Track foot mat scan.

## Results

The demographic and diabetic profile for all participants is shown in Table 1. The kinetic profile at the ankle joint is presented in Table 2, whereas Table 4, shows the clinical implications of ankle joint kinetics for association and prediction of foot ulceration.

**Table 1.** Demographic and diabetic profile of all participants.

| Demographics and Diabetic Profiles | Participants (n=38) (Mean±S.D) |
| --- | --- |
| Age in years | 57.43±4.19 |
| Body Mass Index (BMI) | 25.29 ±2.13 |
| Duration of Diabetes in years | 6±4 |
| Glycated Hemoglobin (HbA1c) in % | 7.1± 1.8 |
| Fasting Blood Glucose (mg/dL) – taken from the most recent lab report | 123±51 |
| Post- Prandial Blood Glucose (mg/dL) – taken from the most recent lab report | 286±94 |

**Table 2.** Ankle joint kinetic analysis of all participants.

| Kinetic Profile variables<br>(Mean±S.D) | Phases of Gait Cycle |  |  |
| --- | --- | --- | --- |
|  | Heel Strike | Mid Stance | Toe Off |
| Peak Plantar Pressure in Kilo Pascals (kPa) | 810±210 | 654±224 | 911±421 |
| Ankle Joint Moment in Newton.meter per kilogram (N.m/Kg) | 1.8±.21 | 1.5±.63 | 2.1±.57 |
| Ankle Power in Watt per kilogram (W/Kg) | 3.1±.81 | 2.9±62 | 3.7±74 |

**Table 3.** Correlation of ankle joint kinetics to risk of foot ulceration (Point-Biserial)

| Kinetic Variables | Correlation<br>r value |
| --- | --- |
| Peak Plantar Pressure in Pascals (Pa) | r= 0.84 |
| Ankle Joint Moment in Newton.meter per kilogram (N.m/Kg) | r=-0.59 |
| Ankle Power in Watt per kilogram (W/Kg) | r=-0.67 |

## Discussion

The present study focused on the kinetic profile among diabetics in the UAE. The ankle joint kinetic was assessed considering that the altered plantar pressure distribution could be the most important etiopathomechanics for the development of diabetic foot ulcers (Pataky et al. 2005, Sawacha et al. 2012, Hazari and Maiya 2020). The altered plantar pressure could contribute to the development of diabetic foot ulcers through distinct mechanisms referred to as Pressure Time Integral (PTI), Peak Plantar Pressure (PPP), and repetitive microtrauma with increased PPP (Hazari and Maiya 2020). The PPP could be the most evident biomechanical change contributing to ulcer formation among diabetics (Hazari and Maiya 2020). In the initial stages, callus formation could be seen with thickened skin, and repetitive stress could break down the ulcer [Yavuz et al. 2008]. The chances of diabetic foot ulcers have been reported to be higher in the neuropathy, however we didn’t assess for DPN. In the present study, we found the mean age and body mass index of participants was 57 years and 25.29 respectively (Table 1), suggesting that the majority of the population was “mid-old” and overweight (Jan and Weir 2021). Understandably, being overweight itself is a factor for increased plantar pressure and the sequels of diabetic changes could have added more to it. The findings of the study suggested that the diabetic population in the United Arab Emirates had increased peak plantar pressure with the highest at the toe-off phase of the gait cycle (Table 1). The meta-analysis conducted by Hazari et al. (2016) based on studies from Rao et al. (2010); Zimny et al. (2004); and Yavuz et al. (2008), suggested that the diabetic population showed significantly higher peak plantar pressure with moderate effects (p=0.03). A recently conducted study by Ahsan et al. (2021) in the Arab region also supported this finding. The study found that the pressure distribution at the heel, midfoot, and metatarsal among diabetics were higher (552.85, 602.10, 482.55 kPa respectively) in comparison to healthy participants (551.25, 527.75, 455.10 respectively). However, in contrast to the study conducted previously, the peak plantar pressure among the diabetics in UAE was considerably high and alarming. A study reported that higher plantar pressure could predispose diabetes mellitus to a higher risk of foot ulceration (Sawacha et al. 2012). Considering this, if we compare the plantar pressure distribution through peak pressure among diabetics in UAE, we could suggest that the risk of ulceration is significantly higher. The means of PPP were significantly higher at all three phases of the gait cycle (810, 654, and 911 kPa) when compared to a population within a similar geographical region as suggested by Ahsan et al. (2021). It should be noted that areas assessed by Ahsan et al. are similar to areas represented through different phases of the gait cycle in the present study and thus comparable. The reasons for higher peak pressure among the diabetic population in UAE need to be assessed through future studies and underlying factors must be determined to avoid the higher risk of ulceration. One of the possible reasons could be the difference in the outcome measures used between the two studies. Nevertheless, it was evident that diabetics in UAE were at higher risk of diabetic foot ulcers with the given findings. In comparison to the previously conducted study by Hazari and Maiya 2020, the range of peak plantar pressure among diabetics in India was 161-266 kPa without and with diabetic peripheral neuropathy. Comparing the present study, the mean value for peak plantar pressure is 3-4 times. The higher risk of foot ulceration among diabetics is also supported by the increased plantar pressure ratio in the forefoot to the hindfoot (Caselli et al. 2002). In the present study, the ratio was found to be 1.12 (911/810 kPa).

Apart from assessing the peak plantar pressure, we also studied the correlational stats between important kinetic variables contributing to the risk of foot ulcerations among diabetics in UAE (Table 2). The risk of foot ulceration was determined by the history of any present or past ulcers, the presence of callus on the plantar aspects of the foot, or any signs of skin breakdown. It was found that the peak plantar pressure has a strong positive correlation, r = 0.84 suggesting that the risk of ulceration was significantly higher. The individual areas of the foot like hallux, metatarsal heads, midfoot, and heel are positively associated with the peak plantar pressure and incidence of foot ulcers (Ledoux et al. 2013). In addition, ankle joint moment and power showed moderate and negative correlation (Table 2) suggesting that reduced ankle joint moment and power could increase the risk of foot ulceration among the UAE population with diabetes mellitus. A reduction in lower limb joint moment and power has been reported among the diabetic population (Hazari and Maiya 2020). The joint moment and power are contributing factors for static and dynamic controls of the joint. Therefore, the significantly increased peak plantar pressure could be attributed to changes in the kinetics at the ankle joint which is directly influenced by the ground reaction force and site of foot ulceration. However, it is understandable that biomechanical changes at one joint would lead to altered biomechanics at subsequent joints in close chain relationships and should be assessed as well.

## Limitations

The present study didn’t compare the ankle joint kinetics of the healthy population to the diabetic population. In addition, the objective of the study was to assess the major kinetics of the ankle joint only where multiple biomechanical factors play a role and could affect the findings of the study.

## Conclusion

The diabetic population in the United Arab Emirates could be at higher risk of foot ulceration with significantly increased peak plantar pressure. The altered kinetics such as reduced ankle joint moment and power could further add to the risk. Underlying factors contributing to the altered kinetics need to be explored.

## Data Availability

All relevant data are within the manuscript and its Supporting Information files.

